# Outcomes and reinterventions following uterine artery embolization for the treatment of uterine leiomyomata

**DOI:** 10.1101/2024.02.03.24302280

**Authors:** Jonathan G. Martin, Alexis M. Medema

## Abstract

**Purpose:** Uterine leiomyomas are common benign tumors that arise from smooth muscle and can significantly impact quality of life. Over the past two decades, uterine artery embolization has risen as a minimally invasive alternative treatment to hysterectomy or myomectomy for the management of leiomyomas. While prior work has established the safety of this procedure, there exist few reports quantifying sequelae, notably rates of subsequent treatment and primary ovarian insufficiency. The purpose of this study is to demonstrate the efficacy of uterine artery embolization as well as to investigate the frequency of gynecologic reintervention and primary ovarian insufficiency following treatment.

**Methods:** The study cohort consisted of patients (n=199) who presented with symptoms concerning for uterine pathology and with leiomyoma(s) confirmed by MRI. This cohort underwent embolization between January 2013 and December 2018 at a single academic institution. Data was collected from retrospective chart review and included demographics, symptomology, imaging, procedural details, and follow-up care. This data was subsequently analyzed to quantify the frequencies of various outcomes at 4–10 years following embolization.

**Results:** Of 199 symptomatic patients with confirmed leiomyomas, all underwent technically successful uterine artery embolization. At the time of follow-up, information was available for 188 (94.5%) patients, of which 145 (77.1%) reported significant symptomatic improvement while 34 required additional intervention—either medical (9%) or surgical (9%). The most common secondary medical management involved hormone therapy, while the most common subsequent gynecologic procedure was a hysterectomy. Additionally, there were seven (3.7%) cases of amenorrhea following embolization.

**Conclusion:** Given its minimally invasive nature, rapid recovery time, and uterine-sparing capability, uterine artery embolization should be considered a frontline therapy for symptomatic leiomyomas. This study supports an overall low complication rate, limited hospitalization time, near-complete resolution of symptoms, and low risk of ovarian dysfunction for a majority of patients. Following embolization, only 9% of patients required additional medical management, and only 9% required a second procedural intervention.

## INTRODUCTION

Uterine leiomyomas, commonly referred to as uterine fibroids, are benign tumors of the female pelvis arising from smooth muscle. With an estimated cumulative incidence that exceeds 80% in African Americans and approaches 70% in Caucasians by age 50^1^, symptomatic leiomyomas are the leading indication for hysterectomy in premenopausal patients and are estimated to cost up to $34.4 billion annually in the U.S^2,3^. For affected patients, leiomyomas are found to significantly impact both physical and emotional quality-of-life (QoL) and are associated with a level of disability similar to that of other chronic diseases^4^.

Uterine leiomyomas most commonly present with heavy menstrual bleeding, and subsequently, chronic iron deficiency anemia requiring iron supplementation or blood transfusion. Patients may also experience a growing pelvic mass, pelvic pain, pelvic pressure, constipation, urinary frequency and urgency, and painful menses^5-7^. Diagnosis is confirmed with ultrasonography, which has the benefit of being widely available and economically accessible, and it may be further characterized with MRI to detail the anatomical environment and arterial supply^5,8,9^.

First described in 1995^10^, uterine artery embolization (UAE) has risen in popularity for the treatment of leiomyomas as an alternative to traditional surgical management. A 1999 outcomes registry established the procedure’s safety, efficacy, and demonstrated symptomatic improvement for a large majority of patients^11,12^. Since that time, further study has continued to support how the minimally invasive nature of UAE promotes a long-term QoL similar to that of hysterectomy or myomectomy, but with shorter hospital stays, shorter time to resumption of normal activity, and lower total costs for hospitals^6,11-16^.

While UAE safety and efficacy are well-established, the rates at which further intervention is required following embolization are less clear. Previous work has demonstrated that greater levels of incomplete infarction are associated with higher likelihoods of requiring a second procedure^17^, but reintervention rates are based largely on the results of two small randomized controlled trials^14,15,18,19^. Additionally, it has been shown that approximately 7% of patients undergoing UAE experience amenorrhea at one year, raising the question of whether inadvertent ischemia may impact ovarian function and therefore the prospect of future fertility^12^. These outcomes are central to the processes of patient counseling and informed consent prior to embolization.

Today, UAE is recommended by the American College of Obstetrics and Gynecology as a minimally invasive interventional procedure for patients desiring uterine preservation. However, given the possibility of needing reintervention and the uncertainty of reproductive outcomes, current guidelines recommend a clinical practice that is individualized to each patient and their respective preferences^7,20,21^. As UAE has grown as a common treatment option for leiomyomas, the purpose of this study is to provide granular data for two common but less analyzed post-procedure sequelae: reintervention and primary ovarian insufficiency (POI) following embolization.

## METHODS

### Data collection

A clinical research and quality improvement query tool was utilized to identify patients who underwent UAE at a single academic institution between January 2013 and December 2018. Data regarding baseline demographics, presenting symptomology, radiographic impressions, procedural details, and reintervention methods were collected from direct retrospective chart review conducted by a single researcher. Outcomes data were collected in 2023 and thus reflect post-embolization timelines ranging from 52 months to 123 months. This study was approved by the Institutional Review Board of <redacted> and the need for individual consent was waived.

### Imaging & procedure

Patients included in this cohort had a leiomyoma diagnosis confirmed by ultrasound and/or MRI. Ultrasound was performed with a transvaginal (TVUS) approach using a wide-band 2-9 MHz linear transducer (GE Healthcare, Boston, MA). Prior to embolization, all patients underwent MRI imaging utilizing a 1.5 Tesla system (Siemens, Munich, Germany).

Embolization of the uterine arteries was conducted by one of six interventional radiologists with one to sixteen years of experience at a single academic institution. Specific technique varied across physicians, but access was typically obtained through either a femoral or radial artery, with the bilateral uterine arteries serving as the baseline target for embolization. Based on either a flush aortogram or on the preprocedural MRI, embolization of one or both ovarian arteries may have also been performed. In general, embolization of each uterine artery was performed to stasis starting with particles sized 500-700μm for up to two vials, increasing to 700-900μm for an additional two vials as needed, and then possibly extending further to 900-1200μm.

### Statistical analysis

Data analysis involved summary statistics, using proportions for categorical variables as well as means and standard deviations for continuous variables. Baseline demographic information included age at embolization and race. Presenting symptoms were classified as abnormal uterine bleeding (including high-volume and high-frequency), bleeding-related chronic iron deficiency anemia, and bulk symptoms (including sensations of pelvic pressure, heaviness, bloating, or urinary symptoms). Patients were further grouped by presenting symptoms as bleeding dominant, bulk dominant, or bleeding and bulk co-dominant. Symptomatic improvement following embolization was determined based on subjective patient report at their follow-up appointments and characterized in a binary fashion as “yes/no” with yes indicating complete or near-complete resolution of symptoms. Outcomes frequencies and risk factors were all analyzed using R version 4.1.3 (Vienna, Austria).

## RESULTS

### Overall outcomes

Initial query identified 199 patients who underwent UAE over a six-year period at a single academic institution. Baseline demographics are presented in Table 1 and revealed an average age at embolization of 44.3 ± 5.3 years. The preponderance of patients were African American (77.9%), followed by Caucasian (14.1%). Upon presentation, the vast majority (93.5%) reported experiencing heavy or otherwise abnormal bleeding, with a significant proportion (72.9%) also describing bulk symptoms. Additionally, 64.8% were found to have chronic iron deficiency anemia on initial laboratory workup.

**Table 1.** Baseline Characteristics.

| Characteristic | (N = 199) |
| --- | --- |
| Age (years) | 44.3 ± 5.3 |
| ≥ 40 years | 167 (83.9%) |
| < 40 years | 32 (16.1%) |
| Race |  |
| African American | 155 (77.9%) |
| Caucasian | 28 (14.1%) |
| Hispanic | 9 (4.5%) |
| Asian | 2 (1.0%) |
| Other | 4 (2.0%) |
| Unknown | 1 (0.5%) |
| Presenting symptoms |  |
| Abnormal Uterine Bleeding | 186 (93.5%) |
| Iron Deficiency Anemia | 129 (64.8%) |
| Bulk | 145 (72.9%) |

Procedural UAE data is summarized in Table 2. The standard approach involved bilateral embolization of the uterine vessels, which occurred in over 96% of cases, though a minority of patients also underwent embolization of either or both ovarian arteries. Typically, the smallest particle size utilized was 500– 700μm. Immediate outcomes revealed an average hospital length of stay of 1.17 ± 1.26 days. There were two patients who experienced prolonged hospitalizations: one who developed *clostridium* sepsis secondary to myonecrosis and was admitted for 17 days, and one who experienced post-procedure pulmonary edema and elevated troponins and was hospitalized for 8 days. There were three additional adverse events in the perioperative period, including one deep vein thrombosis, one radial artery dissection, and one emergency room visit for delayed pain. Altogether, this study demonstrated a periprocedural complication rate of 2.51%. By the time of their respective follow-up, 145 (77.1%) patients reported near-complete symptom improvement following embolization.

**Table 2.** Procedure details.

| Characteristic | (N = 199) |
| --- | --- |
| Length of stay (days) | 1.17 ± 1.26 |
| Periprocedural complication | 5 (2.51%) |
| Artery embolization |  |
| Bilateral uterine arteries | 192 (96.5%) |
| Unilateral uterine artery | 6 (3.0%) |
| Bilateral ovarian arteries | 3 (1.5%) |
| Unilateral ovarian artery | 11 (5.5%) |
| Smallest embolization particle (µm) |  |
| 500 – 700 | 190 (95.5%) |
| 300 – 500 | 8 (4.0%) |
| 150 – 250 | 1 (0.5%) |

### Reintervention

Of the original cohort of 199 patients, follow-up information was available for 188. Overall outcomes are presented in Table 3. Medical therapy alone for additional symptom management was provided for 17 (9.04%) patients and was largely comprised of hormone-associated therapies, including GnRH agonists (2.66%), progestins (2.66%), levonorgestrel intrauterine devices (2.13%), and oral contraceptives (1.60%). Also considered within this category were iron supplementation for ongoing anemia (0.53%) and tranexamic acid for recurrent bleeding (0.53%).

**Table 3.** Outcomes following UAE.

| Outcome | (N = 188) |
| --- | --- |
| Near-complete symptom improvement | 145 (77.1%) |
| Medical reintervention | 17 (9.04%) |
| GnRH agonist | 5 (2.66%) |
| Progestin | 5 (2.66%) |
| Levonorgestrel intrauterine device | 4 (2.13%) |
| Oral contraceptive | 3 (1.60%) |
| Iron supplementation | 1 (0.53%) |
| Tranexamic acid | 1 (0.53%) |
| Procedural reintervention | 17 (9.04%) |
| Hysterectomy | 13 (6.91%) |
| Endometrial ablation | 2 (1.06%) |
| Myomectomy | 2 (1.06%) |
| Amenorrhea | 7 (3.72%) |
| Age ≥ 40yo (N = 156) | 6 (3.85%) |
| Age < 40yo (N = 32) | 1 (3.13%) |

In total, 17 (9.04%) patients underwent further invasive gynecologic intervention due to ongoing symptoms. Of these, 15 procedures occurred within 50 months following embolization, and there were no clear inflection points suggesting high-risk periods for reintervention (Figure 1). The most common procedure was hysterectomy, with 13 patients (6.91%) undergoing either abdominal, laparoscopic, or vaginal hysterectomy with or without salpingectomy in the follow-up interval. Two (1.06%) patients were treated with endometrial ablation and two (1.06%) patients with myomectomy. Relative to the incident uterine artery embolization, hysterectomy was performed an average of 887 days later; the earliest was performed on post-UAE day 414 and the latest on day 1,967. The two ablations were performed on day 359 and day 1,325, and the two myomectomies were performed on day 65 and day 1,185. All 17 patients who underwent a subsequent intervention initially presented with abnormal uterine bleeding, and 14 (82.4%) presented with iron deficiency anemia. However, there was insufficient statistical evidence to suggest that race, age, or presenting symptoms were risk factors for requiring another procedure (Supplemental Table 1).

**Figure 1.**
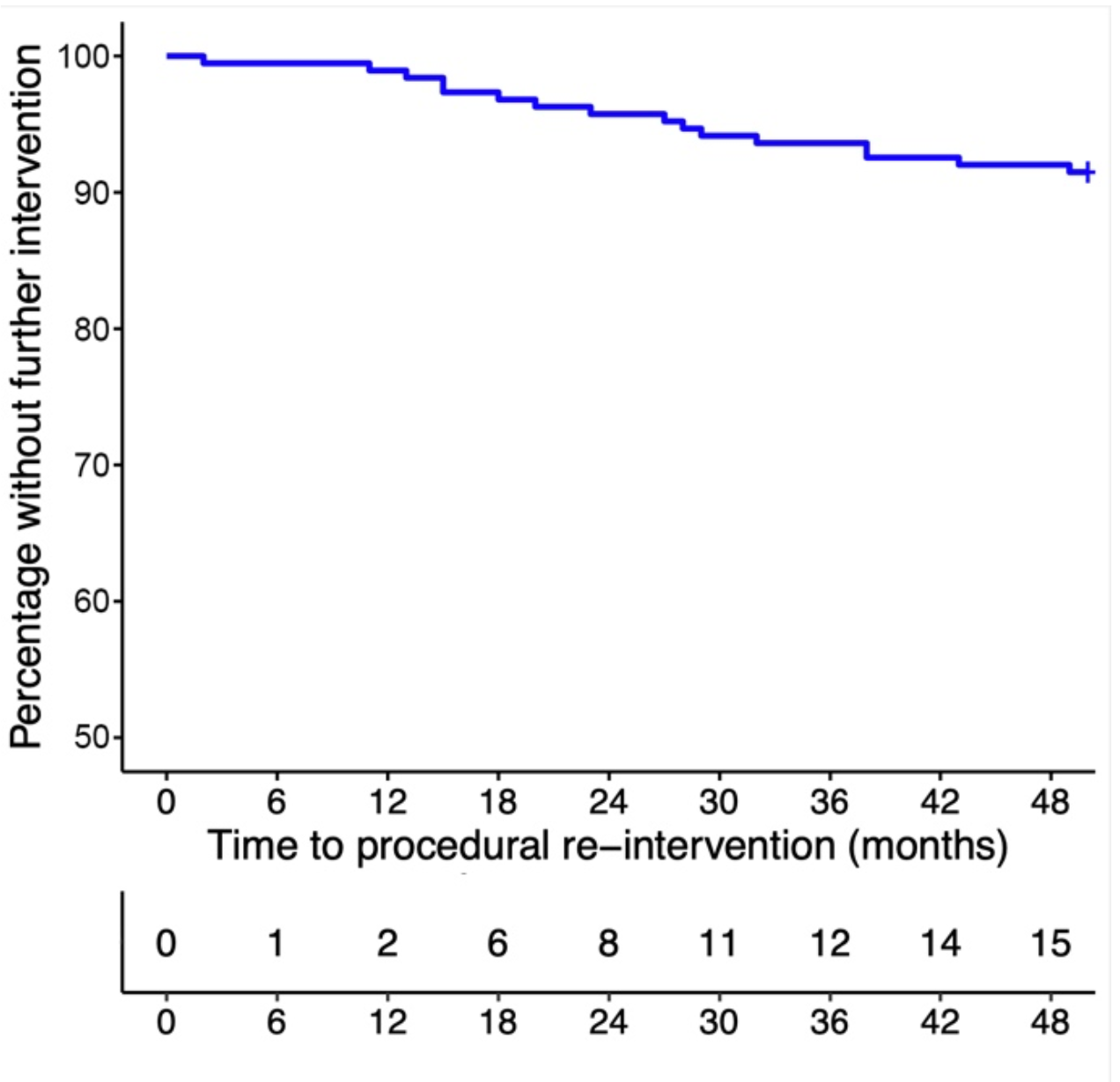
Kaplan-Meier curve demonstrating freedom from procedural reintervention following UAE.

### Primary ovarian insufficiency

Onset of amenorrhea was the main measure of the impact of embolization on ovarian function in this cohort. Of those completing follow-up, seven (3.72%) reported amenorrhea. Strictly defined, primary ovarian insufficiency is ovarian dysfunction occurring before the age of 40. In this cohort, there were 32 patients younger than age 40 at the time of embolization. Among this group, there was one case of post-procedural amenorrhea, suggesting a primary ovarian insufficiency rate of 3.13%.

## DISCUSSION

Since its advent in 1995^10^, UAE has risen as an appealing treatment technique for uterine leiomyomas. While hysterectomy continues to provide the most definitive management, the number of hysterectomies performed annually in the U.S. has decreased steadily in recent years, corresponding with increasing rates of UAE^3^. When comparing outcomes between UAE and hysterectomy, as well as between UAE and myomectomy, all treatment methods have demonstrated effective symptom improvement for affected patients and no statistically significant differences in QoL at long-term follow-up^15,16,22^.

In addition to efficacy, the minimally invasive approach of UAE directly promotes a limited admission time and prompts faster resumption to normal activity. This advantage is demonstrated in this study’s average length of stay of 1.17 days, which aligns with findings of previous work^16,23^. Overall technical outcomes were positive, and only 5 patients (2.51%) experienced a periprocedural complication. Beyond the initial admission, 77.1% reported near-complete symptom improvement at follow-up.

Following their procedure, 18% of patients in this study required additional therapy—either medical management or procedural reintervention. Medical reintervention ranged in severity from iron supplementation for the treatment of persistent anemia to placement of a levonorgestrel intrauterine device for additional management of menorrhagia. These methods are both ubiquitous in practice and effective in cost for treatment of abnormal uterine bleeding for patients without leiomyomas. Therefore, these cases reflect how UAE may provide patients sufficient improvement in QoL to the point where residual symptoms are manageable with conservative methods.

Prior to the advent of UAE, leiomyoma management was accomplished primarily through myomectomy or hysterectomy^10^, and in the event of recurring symptoms following embolization, hysterectomy in particular remains an option for definitive management. This study found that 9.04% of patients required an additional gynecologic procedure after embolization through the follow-up period, with the majority (6.91%) of these patients undergoing subsequent hysterectomy. At one year, there was a procedural reintervention rate of 1.1%, representing one myomectomy and one endometrial ablation. In comparison, one randomized controlled trial published in 2007 found an overall procedural reintervention rate of 7.5% and a hysterectomy rate of 3.8% at one year^16^, while another registry published in 2005 found an overall procedural reintervention rate of 9.5% and a hysterectomy rate of 2.9% at one year^12^. That the current study has a lower overall likelihood of a patient requiring a subsequent gynecologic intervention, notably that of hysterectomy, may reflect a summation of interval improvements in imaging, devices, and technique resulting in improved outcomes.

The impact of embolization on ovarian function requires further investigation, as unintended embolization of the ovarian arteries may compromise endocrine function. The term “primary ovarian insufficiency” describes a continuum of disordered menses—encompassing amenorrhea, oligomenorrhea, and polymenorrhea—that reflects impaired ovarian function and presents in a patient under the age of 40^24^. In this study, seven patients reported amenorrhea at follow-up, and one of those patients was aged younger than 40 years at the time of embolization. This study did not evaluate whether amenorrhea was secondary to embolization or menopause, thus these results suggest but cannot confirm a diagnosis of POI. Yet, an overall amenorrhea rate of 3.72% and amenorrhea rate of 3.13% in patients at risk for POI reflects a general preservation of ovarian function. These findings align with a recent meta-analysis that found no effect of UAE on ovarian reserve^25^ and support the hypothesis that the lower rates of successful pregnancy seen in UAE patients relative to myomectomy patients^26^ may be due to an unfavorable endometrial environment as opposed to ovarian dysfunction.

In summary, UAE for the treatment of uterine leiomyomas leads to substantial symptom improvement for the majority of patients undergoing the procedure. However, a minority will eventually require additional medication or a second procedure for further management. An even smaller proportion may be amenorrheic after their procedure, possibly impacting future reproductive outcomes. With UAE continuing to rise in popularity as a minimally invasive, uterine-sparing option for the treatment of leiomyomas, these outcomes are central to discussions with patients regarding choice of therapy and informed consent. As the American College of Obstetrics and Gynecology recommends shared decision-making on the treatments available given a patient’s individual values and preferences^20^, this study seeks to add granular data clarifying the likelihood of common but understudied UAE outcomes.

### Limitations

The limitations of this study include its retrospective nature reflecting the outcomes from a single institution. The presenting symptoms and follow-up resolution of symptoms were collected from chart review and are reliant on patient self-report, and thus they may be prone to bias. Furthermore, symptom outcomes data was reported on a binary basis of “near-complete resolution,” which may fail to capture mild, moderate, or significant improvement that does not quite approach “near-complete” in description. Regarding other outcomes, the reasons for reintervention, both medical and procedural, were not recorded. Similarly, the causative nature of post-procedural amenorrhea was not explored in greater detail. Analysis of the specific indications prompting secondary intervention would help identify whether reintervention was a sequela of embolization or some other cause. Finally, these outcomes were captured within a period ranging from 4–10 years and thus do not reflect a standardized window that may be more predictive of outcomes.

## CONCLUSIONS

Uterine leiomyomata pose significant barriers to quality of life for affected patients. Since its inception in 1995, UAE has risen as a frontline therapy for management. The benefits of this technique include a minimally invasive approach, short hospitalization length of stay, low complication rate, and near-complete resolution of symptoms for the majority of treated patients with low risk to ovarian function.

## Data Availability

All data produced in the present study are available upon reasonable request to the authors

## SUPPLEMENTAL FIGURES

**Supplemental Table 1.** Risk factors for procedural reintervention

| 95% Confidence Interval |  |  |  |  |
| --- | --- | --- | --- | --- |
|  | OR | Lower limit | Upper limit | p-value |
| **Race** |  |  |  |  |
| Black | 1.053 | 0.275 | 3.225 | 0.963 |
| White | 2.643 | 0.500 | 65.77 | 0.275 |
| **Age** |  |  |  |  |
| ≥ 45 years | 1.699 | 0.614 | 4.969 | 0.293 |
| < 45 years | 0.589 | 0.201 | 1.630 | 0.293 |
| **Symptoms** |  |  |  |  |
| Bleeding dominant | 1.134 | 0.378 | 2.998 | 0.826 |
| Bulk dominant | 0.881 | 0.333 | 2.645 | 0.826 |
| Bleeding & bulk co-dominant | 0.422 | 0.146 | 1.333 | 0.107 |

